# Revisional augmentation of residual neuromusculature and training facilitate embodiment and control of a bionic knee prosthesis

**DOI:** 10.64898/2026.08.24.26343866

**Authors:** Tony Shu, John McCullough, Francesca Riccio-Ackerman, Junqing Qiao, Christian Landis, Yanmei Tie, Laura Rigolo, Matthew J. Carty, Corey L. Sullivan, Ged Weischhoff, Paris Myers, Christopher Shallal, Daniel Levine, Seong Ho Yeon, Ethan Chun, Michael Nawrot, Matthew Carney, Hugh M. Herr

## Abstract

Conventional transfemoral amputation disrupts native neuromuscular pathways, limiting prosthetic joint control, sensory feedback, and the perception of the prosthesis as part of the body. To ameliorate these pathologies, we restored the agonist-antagonist relationship of residual muscles in two individuals with above-knee amputation through an interventional surgical revision. Participants trained with a bionic knee prosthesis before and after the surgical revision while generating neuromuscular, cortical, functional, and affective data. Both individuals demonstrated improvements after the revision that could not readily be attributed to training effects, including: 1) increased proprioceptive afferents and stronger activation in cortical regions associated with sensorimotor integration of their missing joints, 2) improved control of the bionic knee during functional tasks including sit-to-stand and stair ascent, and 3) generally greater prosthesis embodiment, proprioception, and phantom limb definition as assessed through questionnaires and interviews. In contrast, training outcomes were more participant-specific and more variably correlated with amount of exposure, especially before the revision. These pilot findings suggest that revisional augmentation of residual neuromuscular tissues to restore agonist-antagonist dynamics may promote sensorimotor coherence and enhance both functional and perceptual integration with a bionic prosthesis, and remaining participant-specific heterogeneities may be attributable to inter-individual difference in residual limbs neuromuscular system, amputation history, and personal beliefs about prosthesis usage.

**One Sentence Summary:** Surgical revision of residual muscles after above-knee amputation can enhance sensorimotor coherence and embodiment of a bionic knee prosthesis.

## INTRODUCTION

Embodiment, the subjective perception of an appendage as an incorporated part of the body, is effortlessly sustained by most individuals while serving a fundamental role in motor control, self-identity, and quality of life. Embodiment can be divided into three conceptual subcategories: body representation (the mental image of one’s body), agency (the sense of control over movements), and ownership (the feeling that an appendage belongs to oneself) (*1–4*). Following major limb amputation, individuals experience a disrupted sense of embodiment (*5*). Subsequently, the extent to which a prosthesis can become embodied may underpin post-amputation satisfaction and acceptance: without a connection to their prosthesis, individuals are more likely to reject or abandon their device (*6–8*).

Intact human physiology depends on afferent feedback generated by proprioceptive mechanoreceptors to achieve precise joint control and sustained embodiment (*1*, *9*). In cases of limb amputation, these interactive muscle-tendon dynamics are disrupted by the transection of soft and hard tissues. Conventional limb amputation surgeries typically prioritize wound healing and repurposing residual soft tissues toward padding for a prosthetic socket (*10*, *11*). Such techniques neglect restoration of the physiological muscle-tendon dynamics that may be essential for achieving deeper levels of prosthesis embodiment (*11*).

Recent surgical innovations and sensory feedback systems have been developed to improve the perception, embodiment, and functional capacity of prosthetic legs (*12–14*). For example, peripheral nerve stimulation has been shown to improve perception, walking speed, embodiment, and reduce phantom limb pain (*15–17*). Improvements to sensation and function have also been demonstrated with closed-loop spinal cord stimulation (*18*), plantar somatosensory restoration (*19*), peripheral nerve cuffs (*20*), and osseointegration (*21*). Sensory feedback has also been shown to increase prosthesis embodiment and reduce the perceived weight of the prosthesis (*22*). However, translating stimulation-based sensory feedback remains challenging because it often necessitates considerations of implanted hardware and long-term reliability.

The agonist-antagonist myoneural interface (AMI) is a surgical construct and digital sensing configuration that specifically aims to re-establish dynamic muscle relationships that are typically disrupted by amputation (*23*). In doing so, the AMI may restore a more physiological sense of proprioception to the user for improved control of residual musculature and bionic prostheses without the strict requirement of implanted hardware or stimulation (*24–29*). Prior neuroimaging studies also suggest that the AMI may preserve proprioceptive sensorimotor neurophysiology (*30*). Sustaining peripheral proprioceptive pathways through implementation of the AMI could also promote neuronal recovery within the brain, resulting in cortical activity more closely resembling that of individuals without amputation (*30*). Although the AMI has been successfully implemented in humans in the acute case for both transtibial and transfemoral amputations (*27*, *28*), its efficacy has yet to be assessed in the within-subject revisional case. Given the preponderance of individuals living with conventional amputation, it is clinically imperative to understand whether these individuals could similarly benefit from the rehabilitative advantages offered by the AMI, and whether structured prosthetic training could further enhance functional and perceptual bodily integration.

Here, we evaluated the extent to which the implementation of the AMI affects neuromuscular control and prosthesis embodiment in two individuals with existing conventional transfemoral amputation. Toward this end, they were observed for changes in afferent neuromuscular signaling and cortical reorganization over the course of pre- and post-operative prosthetic training periods. Changes in coordination and functional motor control were also evaluated through tasks including free space movement, sit-to-stand transitions, and stair navigation. Their subjective experiences of prosthesis embodiment were evaluated through concurrent questionnaires and structured interviews. We hypothesized revisional implementation of the AMI would produce measurable physiological, psychological, and functional recovery independent of any changes attributable to repeated bionic prosthetic training.

## RESULTS

### Revision of residual transfemoral musculature into agonist-antagonist pairs

Two individuals with prior unilateral conventional transfemoral amputation were recruited with informed consent for this study (Table S1). After completing all experimental tasks during the pre-operative testing week (Table S2), each participant underwent surgical revision to construct an AMI muscle pair for knee flexion and extension in their residual limb (Fig. 1). In both individuals, the AMI was formed using the residual biceps femoris and rectus femoris muscles which had been previously shortened from their physiological lengths due to the original amputation (Supplementary Methods). Additional participant-specific surgical revisions were performed in both participants at the time of intervention to prophylactically prevent the formation of symptomatic neuromas and partially restore the antagonistic relationships between ankle dorsiflexion and plantarflexion (Fig. S1).

**Fig. 1.**
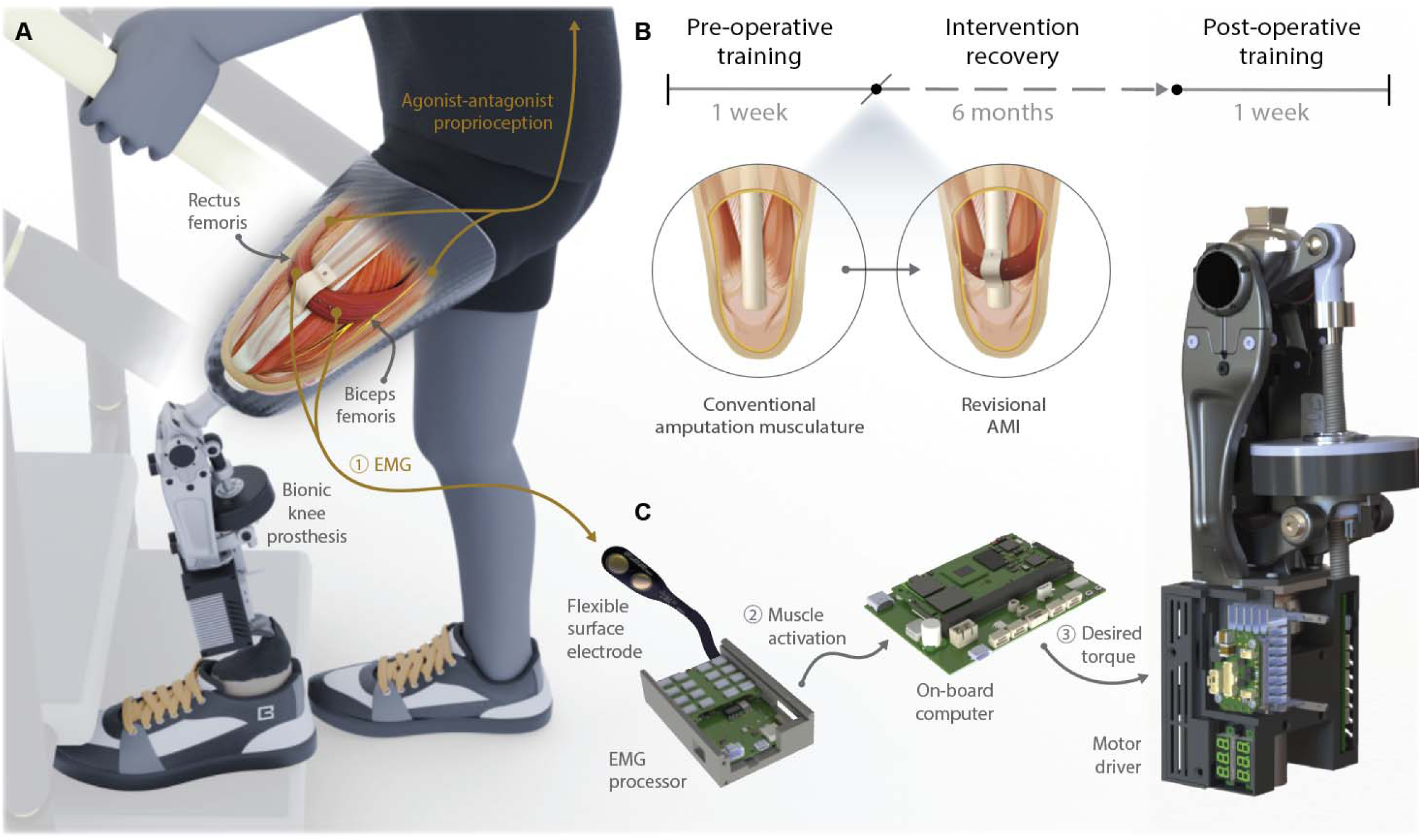
Bionic prosthesis control scheme and experimental timeline. (**A**) Stylized cutaway representation of the revisional agonist-antagonist myoneural interface (AMI) implemented in the residual limb of a person with transfemoral amputation. The transfemoral AMI, encompassing neuromuscular constructs and artificial sensor components, is enclosed in a conventional prosthetic socket to which the bionic knee prosthesis is attached. (**B**) Experimental timeline undertaken by pilot participants (*n* = 2) to determine outcomes associated with the surgical implementation of a revisional AMI in the context of training with a bionic knee prosthesis. (**C**) Electronics mounted externally on the bionic knee prosthesis and socket detect within-socket surface electromyography (EMG) to compute a desired torque at the prosthetic knee joint.

### Surgical revision to create agonist-antagonist relationships augments peripheral neuromuscular function

Evidence from studies on cohorts with transtibial amputation suggests that the quality and amount of afferent proprioceptive feedback from the residual limb directly correlates with motor control accuracy, bandwidth, and bilateral coordination of the corresponding phantom joints (*25*, *26*, *31*). Here, residual knee muscle function was assessed from measurements of graded fascicle strain and muscle activation before and after surgical revision to implement the transfemoral AMI (Fig. 2A and Movie S1). These data were combined to estimate the net amount of proprioceptive information returned to the central nervous system from Type II muscle spindles using an established methodology (*27*, *28*). Agonist muscle contraction and antagonist muscle strain were observed to increase in magnitude post-operatively (Fig. 2B). Correspondingly, both individuals produced greater maximum net afferent firing rates with increased gradation in both muscles post-operatively (*P* < 0.001; Fig. 2C). These effects can be strongly considered to be independent of exposure to training with the bionic prosthesis due to relevant data collection being performed at the beginning of each training week (Table S2).

**Fig. 2.**
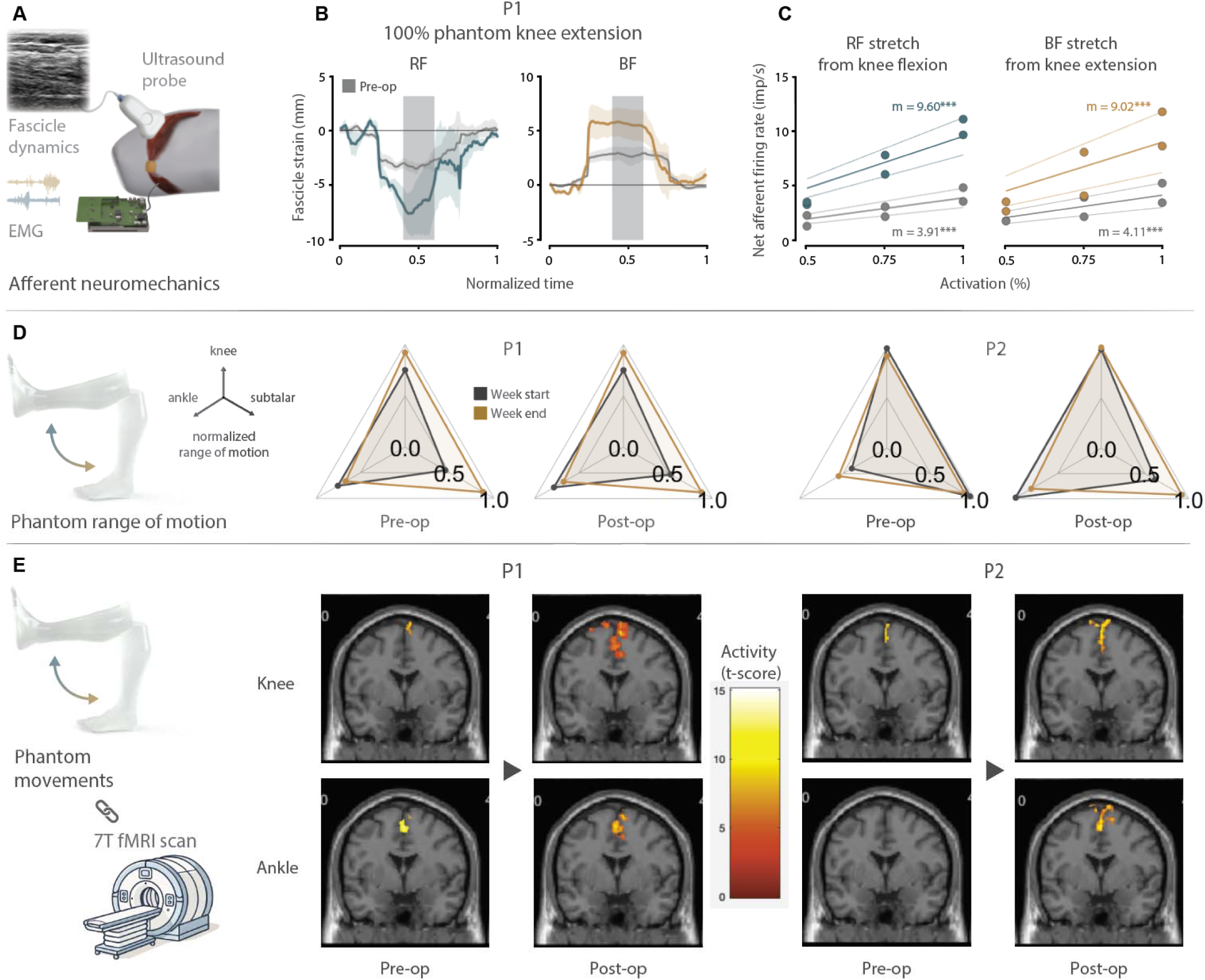
Physiological measures altered following agonist-antagonist revision. (**A**) Measurement of muscle fascicle state and activation from ultrasound and EMG to estimate afferent proprioceptive feedback. (**B**) Representative fascicle dynamics from Participant P1 during phantom knee extension at maximum effort, measured pre- and post-operatively (*n* = 3 trials per trace). Gray, pre-op; teal, post-op rectus femoris (RF) strain; orange, post-op biceps femoris (BF) strain. Gray box, region of steady-state exertion. Shaded regions, SEM. (**C**) Aggregate net afferent firing rates for Participants P1 and P2 for each antagonist muscle at three levels of activation effort, measured pre- and post-operatively. Average net afferent firing rates calculated at steady-state (*n* = 3 trials per marker). Slopes (*m*) are reported (*n* = 6 samples per linear regression, \*\*\**P* < 0.001). (**D**) Phantom range of motion (ROM) assessed through mirrored joint movement for both individuals, measured pre- and post-operatively. Assessments were performed at the beginning and end of the training week (Table S2). (**E**) Cortical activity during performance of phantom knee and ankle flexion and extension as estimated through functional magnetic resonance imaging (fMRI) analysis, measured at the end of pre- and post-operative training weeks. Representative coronal plane slices at the same depth show relative activity changes for the same phantom movement task.

In contrast to the direct correlation between AMI revision and augmented proprioceptive feedback, perceived phantom joint range of motio (ROM) was observed to vary more with either training or the surgical intervention depending on both the individual and specific joint measured (Fig. 2D). Throughout both the pre-operative and post-operative training weeks, Participant P1 perceived increases in the ROM of both phantom knee and subtalar joints, but improvements at the end of pre-operative training were not retained at the beginning of the post-operative training. When comparing Participant P1’s ROM measured at the beginning of pre- and post-op training weeks, no meaningful changes were observed for any phantom joints attributable to the surgical intervention itself. In the case of Participant P2, ROM of the phantom knee matched that of the unaffected knee in all conditions whereas the phantom ankle demonstrated a large increase in perceived ROM as a direct result of the surgical intervention. As with Participant P1, Participant P2 also demonstrated limited improvements to perceived ROM of the phantom subtalar as a result of post-operative training. A larger increase in phantom ankle ROM was observed of Participant P2 as a direct result of the surgical intervention.

### Revisional augmentation of peripheral neuromuscular function normalizes central sensorimotor activity

Blood oxygenation level-dependent (BOLD) responses of the brain were processed from time-series functional magnetic resonance imaging (fMRI) data collected while participants performed isolated, full ROM phantom joint movements to quantify the cortical resources recruited for motor planning and control (Fig. 2E, Tables S3 and S4). Imaging scans for analysis were performed at the end of each training week using a 7 T scanner. Post-operatively, both participants showed increased activation volumes and higher peak t-values in regions corresponding to the premotor and motor cortices within the paracentral lobule and precentral gyrus, as well as the somatosensory cortex in the postcentral gyrus. This pattern of activation closely resembles the typical physiological engagement observed during movements of the contralateral intact joints (*32*, *33*).

### Revisional augmentation of peripheral neuromuscular function improves prosthetic control

Participants were further assessed on their ability to interact with virtual and physical bionic devices to quantify the influence of the AMI surgical revision on efferent prosthetic control. All experiments were administered both pre- and post-operatively. An experiment encompassing graded phantom knee flexion and extension was administered near the beginning of each training week to determine the ability of each participant to achieve and maintain fixed levels of muscle activation (Fig. 3A). Intended phantom knee movement in terms of direction and normalized effort levels was computed in real-time from multiple channels of recorded surface electromyography (EMG) using the nonlinear orthogonal decomposition (NOD) algorithm (*34*). A graphical user interface provided visual feedback on the current state of their phantom knee movement while a series of target movement directions and effort levels were presented in a pseudorandom order through visual markers (Movie S2). Post-operatively, participants were able to achieve improved grading of intended phantom knee direction activation with less off-target co-activation (Fig. 3B), whereas the relationship between the two variables is pathologically and inversely correlated in the pre-operative condition (Fig. 3C).

**Fig. 3.**
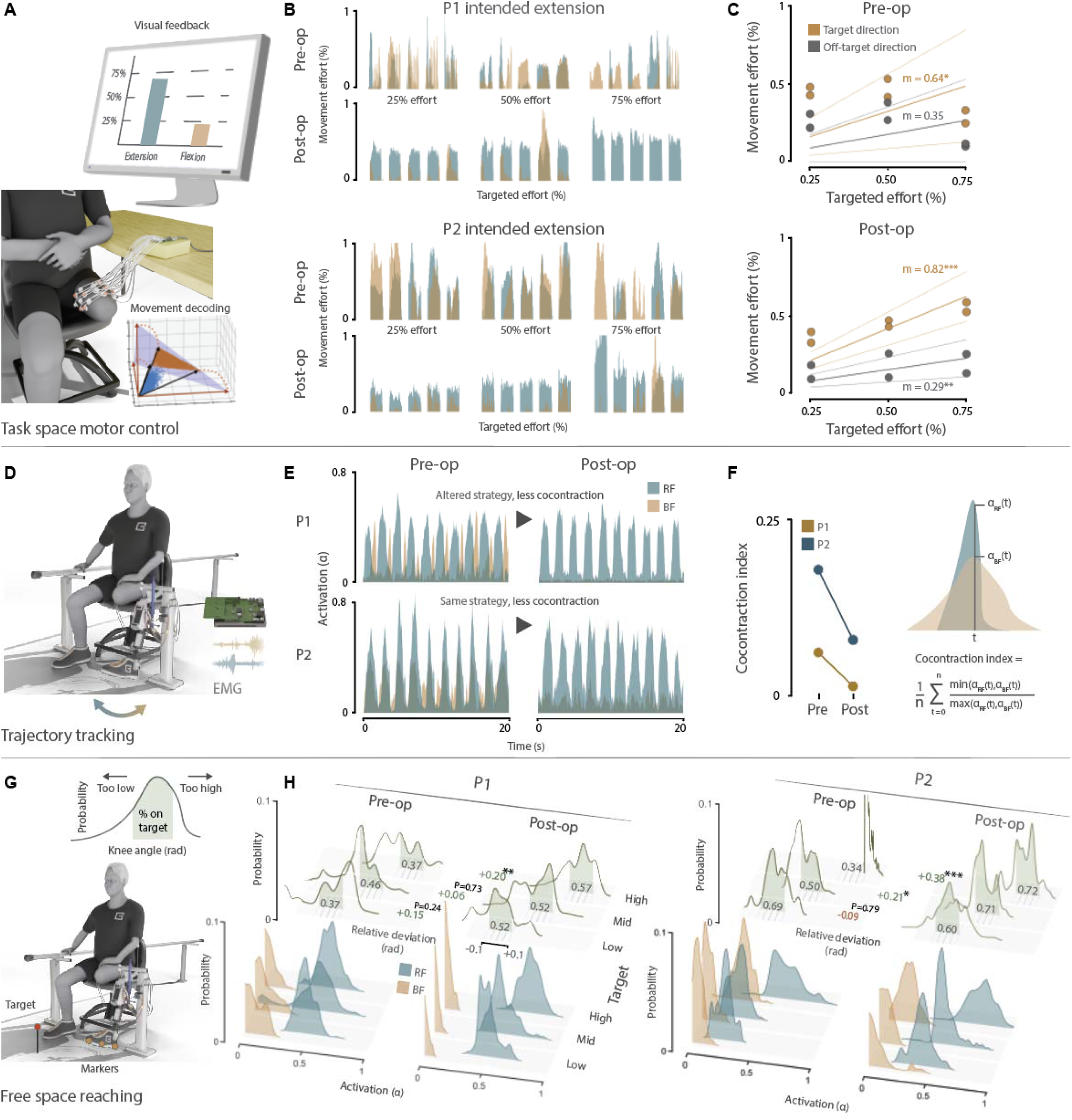
Neuromuscular control improved in free space following agonist-antagonist revision. (**A**) Task space motor control of phantom knee movement with visual feedback. (**B**) Phantom knee extension and flexion efforts extracted from the NOD algorithm at increasing levels of knee extension effort, measured pre- and post-operatively. Blue, extension. Orange, flexion. (**C**) Aggregate on- and off-target efforts during phantom knee extension and flexion at three levels of targeted effort, measured pre- and post-operatively. Average efforts calculated during movement steady-state (*n* = 5 extension and *n* = 5 flexion trials per marker). Slopes (*m*) are reported (*n* = 6 samples per linear regression, \**P* < 0.05, \*\**P* < 0.01, \*\*\**P* < 0.001). (**D**) Trajectory tracking experiment wherein participants mirroring prosthesis motion with their own phantom knee movements. (**E**) Muscle activations at dynamic equilibrium during the trajectory tracking experiment, measured both pre- and post-operatively. (**F**) Co-contraction index indicating the average ratio of lesser muscle activation to greater muscle activation at dynamic equilibrium, measured both pre- and post-operatively. ɑ, normalized muscle activation. RF, rectus femoris. BF, biceps femoris. (**G**) Free space reaching experiment with visual feedback to align markers on the shoe with a fixed environmental target. (**H**) Empirical probability distributions showing each participant’s corresponding knee position and muscle activations at steady-state when reaching for each target, measured pre- and post-operatively. Green shaded regions denote on-target knee positions and changes in accuracy are indicated between pre- and post-operative performance (P1 and P2: *n =* 10, 12, and 12 paired movement accuracies for low, mid, and high targets, respectively [Table S7]; \**P* < 0.05, \*\**P* < 0.01, \*\*\**P* < 0.001).

Participants were also evaluated in a knee trajectory mirroring experiment at the beginning of the week. They were seated and instructed to exercise their phantom knee joint to follow pre-programmed flexion and extension cycles generated by the bionic knee prosthesis (Fig. 3D). The prosthesis was mounted onto the prosthetic socket with thin, flexible electrodes placed within the socket liner to record EMG signals generated by the AMI. Pre-operatively, Participant P1 demonstrated alternating flexor and extensor activations corresponding to movements of the prosthetic knee (Fig. 3E). This pattern evolved post-operatively into one that only contained rising and falling extensor activations in time with knee extension and flexion. In contrast, Participant P2 used the same strategy of rising and falling extensor activations pre- and post-operatively. However, their post-operative activation patterns revealed markedly less flexor co-contraction during extensor activation. Both individuals converged upon a homogenized pattern of muscle activity post-operatively that demonstrated only extensor activation with reduced levels of flexor co-contraction post-operatively (Fig. 3F).

Subsequent experiments investigated myoelectric control of the bionic knee prosthesis across tasks of increasing environmental complexity and user-prosthesis interaction. The first of these assessed each participant’s ability to reach and maintain fixed knee extension angles in free space. Participants were seated adjacent to the bionic knee prosthesis that was mounted in a test fixture such that the shank occupied the anatomical volume of their missing limb while remaining mechanically isolated from their body (Fig. 3G). Participants were provided impedance-based control of knee extension torque determined by graded activation of their extensor muscle while flexor activation was recorded, but otherwise omitted in the control paradigm. Using three markers spaced evenly along the medial aspect of the prosthetic foot, participants were instructed to extend the bionic knee to align each marker with a visual reference on the floor, corresponding to low, middle, and high knee extension targets (Movie S3). Targeted knee extend-and-hold movements progressing from low to high targets were performed in sync with a pre-recorded click track. Compared to their pre-operative performances, both individuals achieved significantly increased targeting accuracy for high targets post-operatively as measured by reduced angular error of the bionic knee joint during the holding period (*P* < 0.01 and *P* < 0.001, respectively; Fig. 3H). Participant P2 additionally demonstrated significantly improved accuracy for the middle target post-operatively (*P* < 0.01). No significant decreases in accuracy were observed in either individual for any target.

A mid-week experiment assessed each participant’s ability to use the bionic knee to perform standing maneuvers from a seated position (Fig. 4A and Movie S4). Ground reaction forces (GRFs) were compared against those generated using each participant’s own prescribed prosthetic device in the same task. For each pair of conditions (operative stage vs. device), participants performed a total of 15 sit-to-stand transitions following a brief practice and tuning period for acclimatization. Torque generated by the bionic knee prosthesis in this experiment was determined from the full calculation of the unified neuromuscular joint controller that considers both flexor and extensor muscle activations (*34*). Bilateral symmetry of normal GRFs was calculated as the cosine similarity between the center of mass of time-normalized GRF trajectories for each leg. For both participants, bilateral symmetry when standing with their prescribed knee prosthesis was not significantly altered post-operatively as they continued to favor their intact leg. For both individuals, using the bionic knee in the pre-operative condition revealed even greater reliance on the intact leg compared to when using their prescribed device. However, post-operatively, both participants achieved significantly improved bilateral symmetry with the bionic knee compared to that obtained by using their prescribed device (*P* < 0.001, both).

**Fig. 4.**
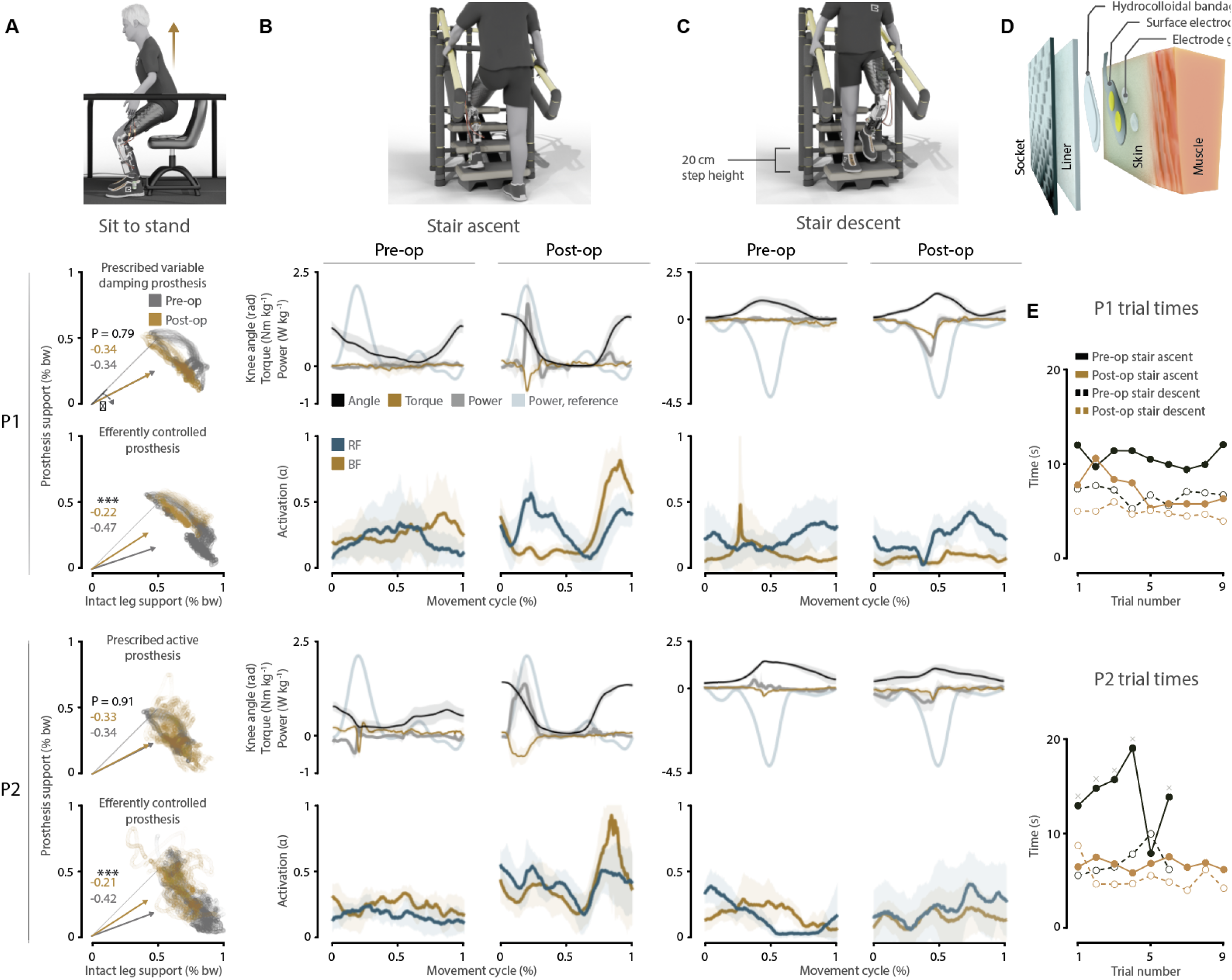
Control of the bionic knee during environmental interactions improved after agonist-antagonist revision. (**A**) Ground reaction forces while standing from a seated position pre- and post-operatively under both conditions. Vectors indicate the centers of mass of aggregated pre-operative and post-operative trajectories. Angular deviations for each vector denote the deflection from perfect symmetry along the X-Y axis. Testing performed for significant differences in mean angular deviation between movements pre- and post-operatively (*n* = 15 paired movements each subfigure [Table S6], \*\*\**P* < 0.001). (**B**) Stair ascent kinematics, kinetics, and muscle activity. Time-normalized trajectories plotted by individual for the affected side during stair ascent performed with the bionic knee, measured pre- and post-operatively (P1: *n* = 18 movements pre-op and post-op; P2: *n* = 12 movements pre-op, *n* = 18 movements post-op). Shaded regions, SEM. (**C**) Stair descent kinematics, kinetics, and muscle activity. Time-normalized trajectories plotted by participant for the affected side during stair descent performed with the bionic knee, measured pre- and post-operatively (P1: *n* = 18 movements pre-op and post-op; P2: *n* = 12 movements pre-op, *n* = 18 movements post-op). Shaded regions, SEM. (**D**) Stylized representation of the sensor configuration used to collect surface EMG from within the socket. (**E**) Time required to ascend or descend the four-step staircase using the bionic knee on the last training day, measured pre- and post-operatively. Each marker represents one attempt. Cross-shaped symbols above a marker indicate the need for a participant to use a pathological step-by-step gait. Missing markers indicate a participant’s voluntary resignation from testing.

Finally, participants were tasked with ascending and descending a set of four steps while using the unified neuromuscular joint controller to modulate torque at the bionic knee (Movie S5) (*34*). Two practice sessions were administered at the beginning and middle of each training week (Table S2). A final data collection session was held at the end of each training week. Each data collection session consisted of nine consecutive trials containing three ascent and descent cycles each. In the pre-operative condition, both participants failed to produce relevant knee extension power during stair ascent and flexion damping during stair descent (Fig. 4B and Fig. 4C). Participant P2 even demonstrated pathological, inverted power curves during stair descent (Fig. 4C). Pre-operative power trajectories during ascent were accompanied by insufficient swing flexion kinematics and muscle activations that generally lacked physiological phase and magnitude. However, post-operatively, both participants demonstrated physiological extension power alongside the emergence of swing flexion kinematics during ascent. Similarly, both participants achieved a measurable degree of flexion damping during descent post-operatively. In these experiments, thin surface electrodes were placed underneath the socket to acquire EMG signals at the surface of the skin (Fig. 4D). These improvements correspond to reduced times required for stair ascent and descent relative to pre-operative durations (Fig. 4E).

### Augmenting peripheral neuromuscular function generally improves embodiment, phantom limb definition, and proprioception

At the beginning of the pre-operative and post-operative testing weeks, participants completed a study-specific questionnaire reflecting on their prescribed prosthesis (Table S2). The questionnaire was designed to evaluate the agency, ownership, and body representation (*2*) of a neurally-controlled powered prosthesis. Of note, this instrument was developed for this study and not a previously validated prosthesis-specific embodiment scale. Each question followed a 5-point Likert scale, ranging from “Never” to “Always,” and responses were correspondingly coded from 1 to 5. Scores for each subcategory were computed by averaging the numeric values across all questions within that category. Participant P2 demonstrated improvements across all three subcategories of embodiment post-operatively compared to pre-operatively (*P* < 0.001 and *P* < 0.01), whereas Participant P1 exhibited no notable change in embodiment of their prescribed device (Fig. 5A).

**Fig. 5.**
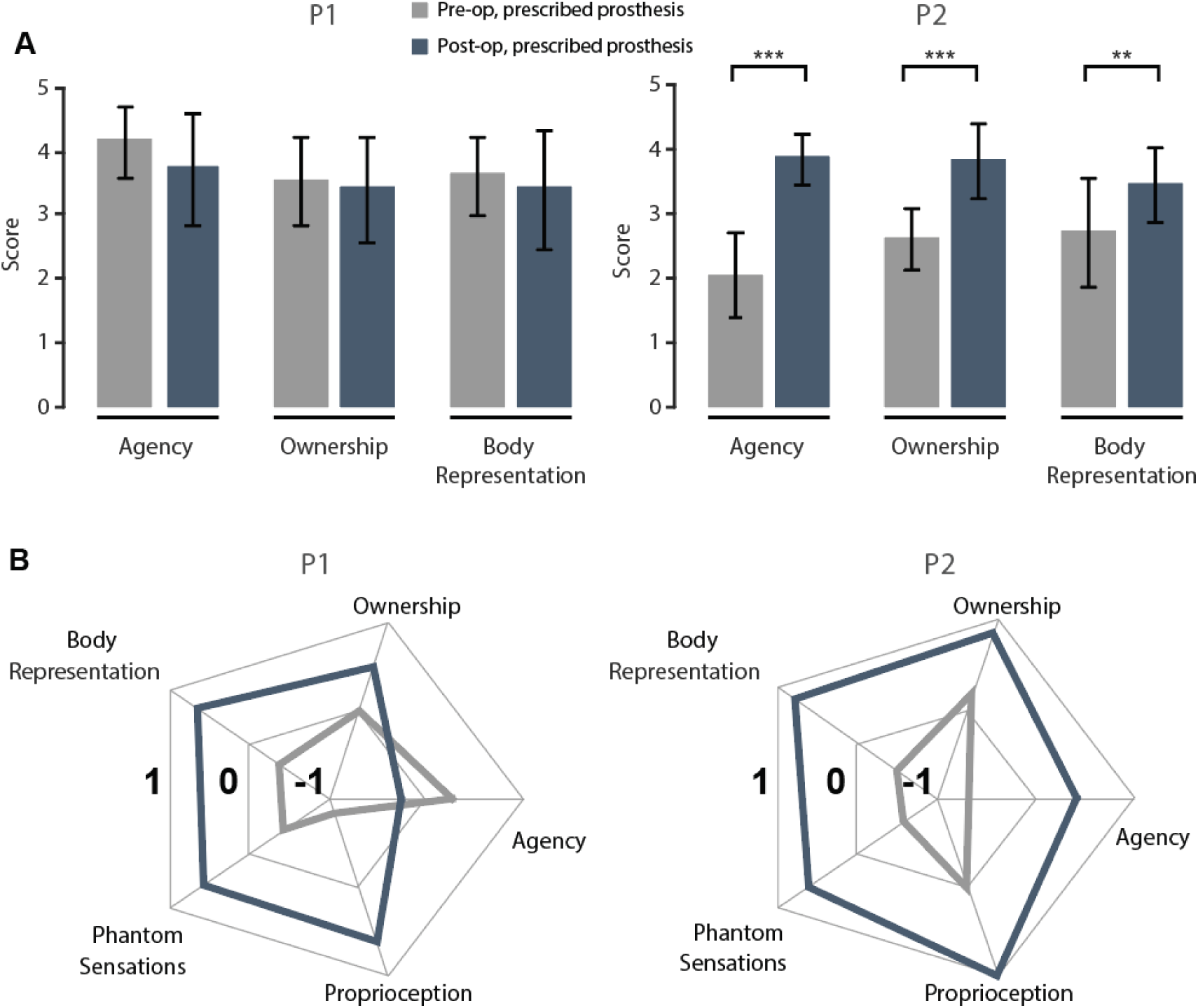
Embodiment of the prescribed prosthesis, phantom limb sensations, and proprioception improved following agonist-antagonist revision. (**A**) Changes in agency, ownership, and body representation, measured using closed-ended questions (*n* = 12 agency questions, *n* = 11 ownership questions, *n* = 14 body representation questions, \*\**P* < 0.01, \*\*\**P* < 0.001). Bars represent mean ± SD. (**B**) Qualitative sentiment changes in embodiment subcategories (agency, ownership, and body representation), as well as phantom sensations and proprioception, derived from thematic coding of semi-structured interviews. A value of 1/-1 means all positive/negative sentiment thoughts for that thematic category.

Semi-structured interviews were conducted alongside the questionnaires to expand on participant responses using sentiment analysis. Transcripts were thematically coded for agency, ownership, body representation, phantom sensations (encompassing phantom pain), and proprioception by a single researcher. Coding was performed at the sentence level, and each sentence referencing a particular embodiment dimension was assigned a sentiment label (positive, neutral, or negative) with a corresponding numeric value (+1, 0, −1). Sentiment scores were computed by averaging the responses within each code category, yielding a normalized sentiment index ranging from −1 to +1. Intercoder reliability was assessed using a second trained coder’s independent labels from three of twelve interviews. Cohen’s kappa and observed agreement were calculated to be 0.75 and 87.7%, respectively, indicating substantial agreement between the two coders in support of the reliability of the sentiment-derived interview analysis (*35*).

Following revisional surgery, both participants reported sentiment responses related to their prescribed prosthesis (Fig. 5B). These trends were reflected in participant narratives. Pre-operatively, Participant P1 described their conventional prosthesis as “inanimate,” saying, “If this were to go away and I’d get a new one, I’d be pretty psyched. I don’t care, you know. It’s a piece of metal.” Post-operatively, the same participant reported, “I can close my eyes and feel my whole leg… my thigh, my knee, down to my calf, to my ankle… I feel like I can visualize all those parts.” Participant P2, who expressed a strong aversive response to phantom pain pre-operatively, “As soon as somebody says the word ‘phantom pain’ I get a tingling feeling in my foot… It triggers something”, described dramatically improved proprioception after surgery, “I can feel my foot on the floor, like literally… I kind of feel like I know where it is at all times now… I didn’t feel it the way I do now before the surgery.”

The deviation from this general post-operative improvement in sentiment of the prescribed prosthesis was a decline of agency for Participant P1. Despite reporting improved socket fit, “The socket that I have now is the best fitting socket I’ve ever had… I do believe that it’s because of the musculature. Now it kind of locks in”, the participant expressed a paradoxical perceived loss of control due to increased awareness of the ability to move the phantom limb: “I don’t feel like I’m there now… I was better at it before. I was less conscious of my abilities before the surgery.”

### Training with the bionic prosthesis improves embodiment, phantom limb definition, and proprioception

After the third and last day of each testing week, participants were administered the same questionnaires on embodiment, phantom limb definition, and proprioception as completed at the beginning of the week (Table S2). During the pre-operative week, Participant P2 demonstrated increased agency and ownership on the last day of bionic training relative to the third day when they were first presented the opportunity to efferently control the bionic prosthesis (*P* < 0.01 and *P* < 0.05, respectively; Fig. 6A). In the post-operative week, Participant P1 demonstrated increased agency and ownership, whereas Participant P2 demonstrated increased agency and body representation (*P* < 0.05, all; Fig. 6B). Semi-structured interviews were conducted at the same time points and analyzed using thematic coding. During the pre-operative week, both participants demonstrated mixed trends, with variable changes across embodiment, proprioception, and phantom sensations (Fig. 6C). However, during the post-operative week, both participants demonstrated small increases in agency, body representation, and phantom sensations with training (Fig. 6D).

**Fig. 6.**
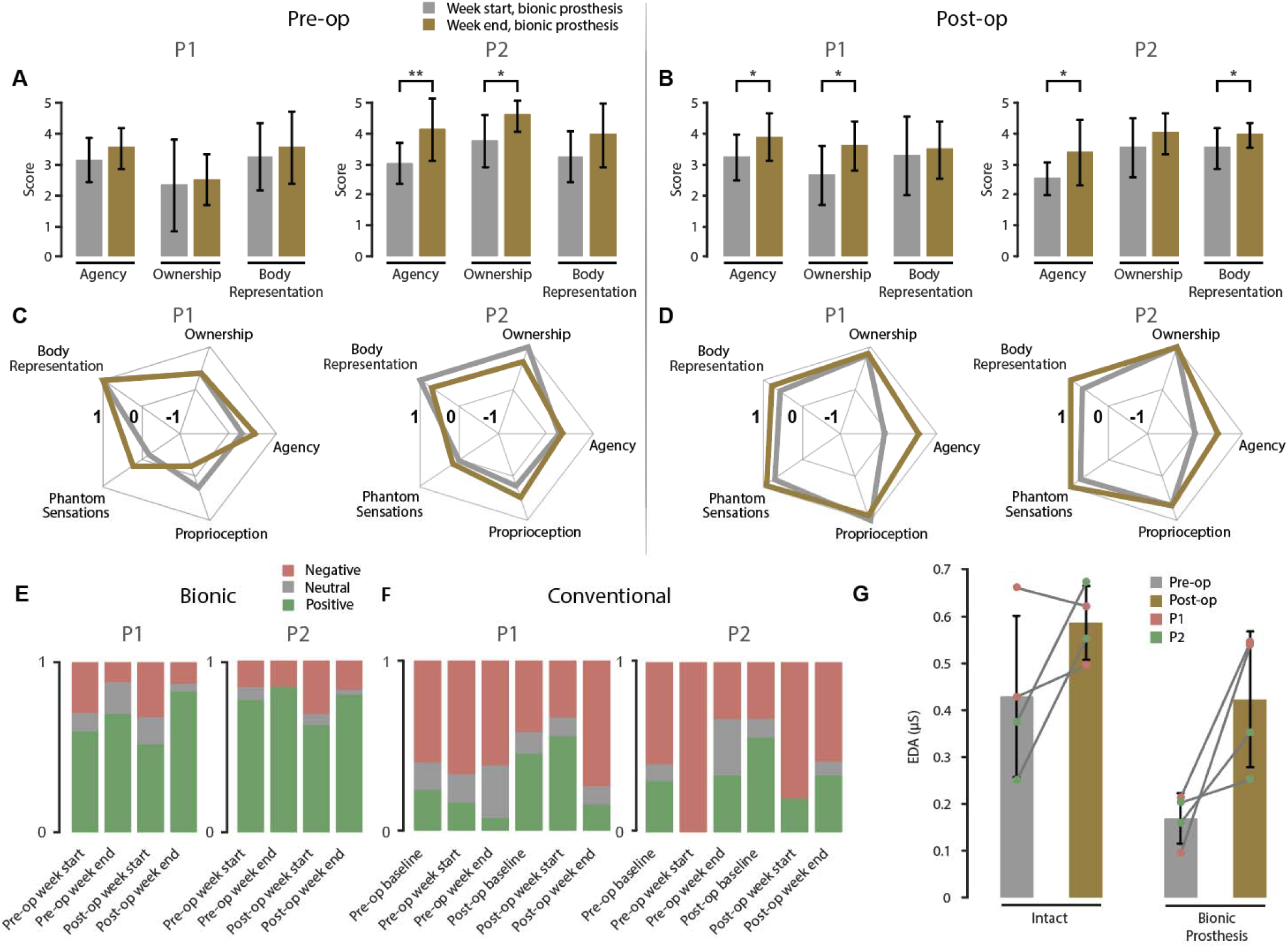
Embodiment of the bionic and conventional prostheses, phantom sensations, and proprioception changed across testing periods. Changes in agency, ownership, and body representation measured using closed-ended questions administered at the beginning and end of the testing week in the (**A**) pre-operative training week and (**B**) post-operative training week (*n* = 12 agency questions, *n* = 11 ownership questions, *n* = 14 body representation questions, \**P* < 0.05, \*\**P* < 0.01). Bars represent mean ± SD. (**C**) Qualitative sentiment changes in embodiment subcategories (agency, ownership, and body representation), as well as phantom sensations and proprioception, derived from thematic coding of semi-structured interviews conducted pre-operatively and (**D**) the same conducted post-operatively. (**E**) Sentiment distribution (positive, neutral, negative) toward the bionic prosthesis, based on coded interview data taken at the start and at the end of the pre-operative and post-operative training weeks. (**F**) Sentiment distribution toward the conventional prosthesis from different time points in both pre-operative and post-operative training weeks. Data are shown for both participants (P1 and P2) and reflect qualitative coding of user-reported statements during semi-structured interviews. (**G**) Average EDA (in µS) is shown for each participant across pre- and post-operative testing sessions. Error bars, group means ± standard deviation. Markers, individual participant responses.

Qualitative data provided further insight into the perceptual and functional changes across the training period. In the pre-operative week, Participant P1 noted, “I looked at [my wife] after I took a couple steps, and I was like, oh my god this is amazing.” By the end of the week, they stated, “It felt like there was a leg underneath a lot of electronics… but my leg was fully intact.” Participant P2 also reported positive impressions, describing the device as enabling recovery of lost function: “It makes me feel like I can actually get back what I lost a long time ago… walking down the street holding my wife’s hand.”

Using the same interview dataset, we also computed an overall sentiment toward each device by pooling statements across all five thematic domains. Thus, the overall sentiment is influenced by the number of coded statements within each domain such that domains that were discussed more frequently have greater contribution. Sentiment toward the bionic prosthesis was predominantly positive across all testing sessions and slightly increased from the beginning to the end of each training week (Fig. 6E).

For both participants, sentiment towards the bionic prosthesis was lower at the start of the post-operative week than at the end of the pre-operative week (Fig. 6E). At the beginning of the post-operative training week, Participant P1 noted initial difficulties in control of the bionic prosthesis, stating, “I always felt like I was moving it. I just don’t always feel like it did exactly what I wanted it to.” After the training at the end of the week, they reported a stronger connection: “When I was power walking, I was completely, you know, unaware of it being anything but my leg… all the muscles in my residual limb were functioning to power that leg, and I could feel it all the way down.” Similarly, Participant P2 started the week struggling with coordination: “That’s the problem with the stairs too, because I’m visualizing in my head going up, putting down and going over… my muscles are too fast, I guess… the muscles in my stump are reacting a lot faster than I can physically get there.” By the end of the week, they reported a substantial improvement: “Everything is just so much better, so much more amazing… [Now] it’s so much easier to control. It makes me feel, you know, so much more whole as a person.”

To complement subjective reports of embodiment, proprioception, and phantom limb definition, electrodermal activity (EDA) was measured at the end of the week as a marker of physiological arousal in response to non-harmful, joint-specific sensory stimulation. During this experiment, participants were seated with the bionic prosthesis while a sharp metal prop was slowly brought within close proximity to their intact and prosthetic knee and ankle joints (Movie S6). EDA was recorded via skin conductance sensors placed on the fingers and processed. Post-operatively, both participants exhibited generally increased EDA responses compared to their pre-operative measurements, in both the intact and prosthetic limbs (Fig. 6G).

### Augmented peripheral neuromuscular function and repeated exposure to the bionic prosthesis alter sentiment toward the conventional prosthesis

Participant sentiment toward their conventional prosthesis remained predominantly negative throughout the study period (Fig. 6F). For Participant P1, pre-operative baseline descriptions emphasized a lack of integration: “It’s like basically attaching a stick to the end of your limb. It’s inanimate. It doesn’t have any interaction.” By the end of the week, they acknowledged familiarity but maintained low sentiment: “It’s okay. It feels fine. You know, I don’t hate it… It feels like a prosthetic. It feels like an aid to my amputated leg.” Post-operatively, this sentiment remained low. At the start of the week, they noted, “The socket felt inanimate… I flex muscles, and nothing happens. It’s completely useless.” At the end of the week, they described the prosthesis as “more like a well made crutch than it does a leg.”

Participant P2 showed a similar pattern of sentiment toward their conventional prosthesis. At the pre-operative baseline assessment, they commented on their recently acquired socket with, “Now that I have this new socket… the leg actually feels more like mine.” During the pre-operative week, they also said, “I appreciated my leg, because I’m used to it.” By the end of the post-operative week, they stated, “It made me sad [to return to the conventional prosthesis]… I don’t hate my conventional, but the difference is its ability to respond back to my flexion.” They concluded, “The conventional… is connected to me, but the bionic moves more like a part of me.”

## DISCUSSION

Two individuals with preexisting unilateral transfemoral amputation received a surgical intervention to revise extant soft tissues into antagonistic AMI muscle constructs. Previous studies have correlated the AMI with increased proprioceptive feedback, improved neuromuscular control, and improved efferent prosthetic control in individuals with transtibial amputation (*25*, *26*, *31*) and transfemoral amputation (*28*). By controlling for the amount of exposure to the bionic prosthesis, our data tentatively propose a causal relationship between implementation of the AMI and observed improvements that include partial recovery of proprioceptive feedback and elevated cortical activity in regions associated with lower-extremity joint control. Our data also suggest the AMI may provide these improvements in a revisional context as opposed to only from an acute surgery, though this claim would benefit from validation in follow-up studies with appropriate sample size to mitigate the influence of individual factors including surgical history and residual limb anatomy.

Questionnaires and interviews at the beginning of each week demonstrated that the AMI revision surgery likely enhanced each participant’s sense of prosthesis embodiment. Improvements were most pronounced in agency and ownership and were supported by participants’ detailed descriptions of phantom joint awareness and control (Fig. 5). Participants also reported improvements in phantom limb sensation and proprioception following surgery (Fig. 5). Prior to the AMI procedure, phantom pain and imprecise phantom awareness were common. However, post-operatively, participants were able to describe the position, shape, and movement of their phantom limbs in greater detail. They reported being able to mentally move phantom joints and feel their toes and feet on the ground. Complementing these subjective reports, EDA responses to non-harmful limb-specific stimuli increased post-operatively in both the intact and prosthetic limbs (Fig. 5). These physiological results suggest heightened sensorimotor engagement and may reflect improved central integration of peripheral afferents.

Implementation of the AMI using knee flexor and extensor muscles may have also augmented control of more distal phantom joints through synergistic muscle activations. As a likely outcome of the surgical intervention alone, both participants demonstrated approximately twice their original maximum net afferent firing rates during phantom knee movements after recovering from surgery and before additional exposure to efferent control of the bionic prosthesis (Fig. 2C). Post-surgical fMRI scans also revealed generally increased peak t-values and greater volumes of activated cortical regions associated with phantom joint movements in both participants compared to pre-operative scans at the same time point (Fig. 2E and Table 1). In particular, these increased levels of cortical activity relative to pre-operative levels persisted during isolated phantom ankle dorsiflexion and plantarflexion despite the individual-specific variations in neuromuscular tissues used to construct the AMI for ankle control. Specifically, Participant P1 received an AMI for ankle control fashioned from two denervated knee flexors reinnervated by the common peroneal and tibial nerves whereas Participant P2 received an AMI for ankle control fashioned from a single denervated knee flexor reinnervated by adjacently inserted tibial and common peroneal nerves (Fig. S1). Although this preliminary evidence further suggests that it is the antagonistic mechanical relationship between muscles rather than original residual neurophysiology that is key to neuromuscular function, a follow-up study involving a larger population of participants with transfemoral amputation and ankle AMI constructs would be required to assess the recovery of distal phantom joint function. Experiments performed should encompass methodologies to assess functional control of distal prosthetic joints, and separately, simultaneous recordings of muscle state and cortical activity to quantify the physiological correlation between peripheral and central nervous system activity. As a final consideration, Participant P2 produced lower overall peak t-values during movements of the phantom knee and ankle (Fig. S3). This heterogeneity may be attributed to Participant P2’s earlier and more traumatic original amputation relative to Participant P1’s elective amputation, resulting in an exacerbated degree of muscle atrophy following already greater soft tissue damage (Table S1).

**Table 1.** Cortical activity observed during phantom joint movements performed at the end of testing weeks.

| **P1** | **Left knee (number of voxels)** |  |  |  | **t<sub>max</sub>** | **Left ankle (number of voxels)** |  |  |  | **t<sub>max</sub>** |
| --- | --- | --- | --- | --- | --- | --- | --- | --- | --- | --- |
|  | Paracentral lobule | Precentral gyrus | Postcentral gyrus | Supplementary Motor Area |  | Paracentral lobule | Precentral gyrus | Postcentral gyrus | Supplementary Motor Area |  |
| Pre-op | 839 | 8 | 197 | 517 | 11.13 | 504 | 6 | 59 | 603 | 7.48 |
| Post-op | 1121 | 72 | 557 | 1621 | 19.08 | 765 | 9 | 52 | 930 | 11.82 |

| **P2** | **Left knee (number of voxels)** |  |  |  | **t<sub>max</sub>** | **Left ankle (number of voxels)** |  |  |  | **t<sub>max</sub>** |
| --- | --- | --- | --- | --- | --- | --- | --- | --- | --- | --- |
|  | Paracentral lobule | Precentral gyrus | Postcentral gyrus | Supplementary Motor Area |  | Paracentral lobule | Precentral gyrus | Postcentral gyrus | Supplementary Motor Area |  |
| Pre-op | 454 | 90 | 48 | 410 | 9.71 | 110 | 96 | 20 | 1 | 5.41 |
| Post-op | 945 | 175 | 156 | 954 | 12.08 | 578 | 134 | 20 | 790 | 7.66 |

Regarding phantom joint perception, our findings tentatively suggest that training with a bionic prosthesis may influence phantom joint ROM whereas surgical revision of peripheral neuromusculature does not necessarily change it (Fig. 2D). The results raise multiple hypotheses. First, a physiological range of phantom knee motion may be attainable with only a modest level of proprioceptive feedback compared to the phantom ankle joint and its associated muscles. This is suggested by the relatively complete and persistent ranges of phantom knee motion demonstrated by both participants pre-operatively before observing the increased net afferent firing rates from the associated knee muscles post-operatively (Fig. 2C). The results are surprising when considering a previous study which found that those with an ankle AMI demonstrated significantly increased range of phantom ankle motion compared to those with conventional transtibial amputation (*26*). However, this is tempered after considering a second study (*27*) with similar populations which found that those with conventional transtibial amputation demonstrated negligible net afferent firing rates, whereas the two individuals of this study already demonstrated slight, but positive net afferent firing rates graded with antagonist stretch pre-operatively (Fig. 2C). Second, training with a bionic knee prosthesis may be able to extend the ROM of adjacent phantom joints independent of underlying peripheral neuromuscular configurations. For Participant P1, phantom subtalar ROM increased throughout pre-operative and post-operative training weeks (Fig. 2D). Participant P2 also demonstrated an increase in phantom subtalar ROM throughout the post-operative training week. Considering how neither participant demonstrated obvious changes to phantom ankle ROM as a result of training, and that phantom subtalar ROM reset to pre-operative values for Participant P1 at the beginning of the post-operative training week, this adjacency effect may be weak and sustained only through continuous bionic training.

Both participants demonstrated improvements in performance during free space control tasks with visual feedback after the surgical intervention and before donning the bionic prosthesis. In the virtual control task with visual feedback, both participants generated estimated movement efforts associated with an intended direction of knee movement that were positively graded with targeted effort percentages, in contrast to their pre-operative performance (Fig. 3C). Simultaneously, pathological coactivations in the opposing movement direction were reduced at each targeted effort percentage. Further, implementation of the revisional knee AMI was observed to correspond with homogenized muscle activations between participants in the trajectory mirroring task without efferent prosthetic control (Fig. 3E). Both participants converged onto an extension-only strategy with low flexor coactivation that corresponds to what might reasonably be expected of an intact knee being repeatedly extended and relaxed while seated. In the task involving targeted reaching of a physical prosthesis with visual feedback, general improvements in knee positioning accuracy were found to be consistent with improved grading of extensor muscle activations and reduced flexor coactivations, even if not all targets were more accurately acquired after the revisional AMI surgery (Fig. 3H). However, this interpretation is tempered by equipment failures that limited attainment of the number of paired samples specified in the original study design, reducing statistical power (Table S7). We recommend that future studies include redundant data collection sessions to account for inter-subject variability and the inherent limitations of small-cohort studies with experimental prostheses. Overall, these positive performance trends proffer that recovery of proprioceptive afferents may provide a controller-agnostic basis for improving physiological control of prostheses.

Metrics associated with functional tasks involving environmental interactions with the bionic knee improved as a result of implementing the transfemoral AMI. In the sit-to-stand task, both participants demonstrated the greatest symmetry of bilateral GRFs observed in any tested condition when modulating bionic knee extension torque with their transfemoral AMI constructs (Fig. 4A). For Participant P1, the reliance on the bionic prosthesis while standing up from a seated position increased significantly from approximately 25% to 39% of body weight from pre- to post-operative conditions (*P* < 0.001). Similarly, Participant P2 significantly increased support on the bionic prosthesis from 27% to 39% of body weight (*P* < 0.001). This performance is remarkable considering that bilateral symmetry of GRFs when using the bionic prosthesis with conventional amputation musculature was worse than in all other tested conditions. Importantly, there was no significant change in reliance on the prescribed prosthesis while standing up after the surgical intervention (remaining approximately 32% of body weight for both participants). Because both a variably damped prosthetic knee and a powered prosthetic knee were represented, this preliminary result suggests that the revisional transfemoral AMI does not impede the capacity to use commercial prosthetic devices.

Regarding use of the bionic prosthesis for stair ascent and descent, both participants’ improved post-operative performances suggest that the transfemoral AMI is able to produce more physiological gait dynamics than conventional residual musculature after a similar amount of training (Fig. 4B and 4C). Mean peak knee extension power during stair ascent increased from negligible values for both individuals to 1.67 and 1.38 W/kg^−1^ for Participant P1 and Participant P2, respectively. These values remain lower than the peak knee power generated by bilaterally intact subjects during stair ascent, measured at approximately 2.3 W/kg^−1^ (*36*). Staircase traversal times were also reduced for both participants post-operatively (Fig. 4E). Comparing performance during the final trial between pre- to post-operative training weeks, Participant P1 reduced staircase ascent time by 48% from 12.06 to 6.31 seconds and staircase descent time by 41% from 6.71 to 3.94 s. Participant P2 reduced staircase ascent time by 56% from 13.86 to 6.15 seconds and staircase descent time by 32% from 6.17 to 4.18 s. The relatively larger reductions in staircase ascent time over stair descent time are reasonably explained by the post-operative emergence of powered knee extension and full ROM swing flexion during ascent (Fig. 4B) compared to the more minor increase in flexion damping during stair descent (Fig. 4C). The tendency for physiological gait dynamics to emerge in persons with above knee amputation upon restoring neuromuscular function is corroborated in the complementary case of providing sensation of foot contact through intraneural sensory feedback (*16*). The persistent lack of physiological levels of flexion damping during stair descent may be partially attributable to handrail usage. However, commercially-available variable damping and powered prosthetic knees have similarly been observed to only generate up to -2 W/kg of flexion damping during stair descent with minimal handrail assistance (*37*), less than half of the -4.5 W/kg of flexion damping generated by intact knees (*36*). It is plausible that the reduced flexion damping of commercially-available prosthetic knees can be attributed to electromechanical design tradeoffs that limit peak power as opposed to modest handrail usage. Future users of our bionic knee may be able to generate physiological levels of flexion damping during stair descent with further training to become accustomed to its greater power limits compared to commercially-available knees. Altogether, both participants’ improved performances during these functional tasks suggest that recovery of proprioceptive afferents may facilitate efferent prosthesis control with capabilities beyond those afforded by conventional prostheses, but efferent prosthesis control without sufficient neuromuscular augmentation may not exceed the ability of conventional prostheses.

Beyond functional outcomes, exposure to the bionic device during short-term training may have improved select measures of prosthesis embodiment. Qualitatively, interviews revealed a perceptual shift from operating an external tool to experiencing prosthetic movement as an extension of the self. Closed-ended questionnaires revealed improvements in specific embodiment subcategories, whereas sentiment analysis of interview data identified small or negligible increases in embodiment with training (Fig. 6). Notably, these gains were distributed across different subcategories for each participant rather than uniformly, possibly due to factors including amputation history, motor learning, and user expectations. Together, these data suggest that short-term training may strengthen unique subcategories of prosthesis embodiment per individual, but not in a strongly predictable manner.

Repeated exposure to the bionic prosthesis may have raised expectations of device performance. After initial exposure to the bionic prosthesis, participants expressed satisfaction with the novelty of simple flexion and extension of the bionic knee through muscle contractions. However, by the start of the post-operative week, this excitement was refined into a desire for more precise and reliable control. Interview data revealed that minor discrepancies between intent and resultant prosthesis movement became increasingly poignant sources of frustration. This general inverse relationship between error rate and prosthesis embodiment is consistent with previous observations of transfemoral AMI-based control of a bionic knee prosthesis (*28*). We hypothesize that with increasing exposure and training, greater bionic capability may be required to maintain initial levels of satisfaction, and any perceived errors in control become relatively more detrimental to sentiment toward the bionic prosthesis.

Exposure to an efferently controlled device also appeared to shift participants’ perceptions of their prescribed prostheses. At the beginning of the pre-operative week, conventional prostheses were described in neutral or accepting terms. However, both individuals began to describe their prescribed prostheses as inadequate following exposure to the bionic limb. Phrases such as “a well-made crutch” replaced earlier descriptions that indicated tolerance. These preliminary observations raise the hypothesis that exposure to efferent prosthetic control may alter what users consider sufficient prosthetic function, not only for bionic prostheses, but in general.

Regarding the embodiment questionnaires, the assessments used in this study departed from widely adopted embodiment protocols in two aspects. First, control questions were omitted from analysis as they confused participants and disrupted the flow of interviews. The mixed-methods design, which paired closed-ended questionnaires with semi-structured interviews, was designed to mitigate superficial responses by allowing participants to contextualize each questionnaire response. Nevertheless, this approach does not fully replace the psychometric role of control items and limits our ability to formally assess response specificity. Second, the questionnaire used a frequency-based response scale rather than the agreement-based scales commonly found in other studies on the rubber hand illusion and prosthesis embodiment. This format captured how often embodiment-related experiences occurred during prosthesis use, but it limits direct comparison to prior work and may affect interpretation of the measured construct. Frequency-based ratings may emphasize recurrence or temporal consistency, whereas agreement-based ratings may better capture strength of endorsement, conviction, or perceived intensity. Accordingly, questionnaire-derived embodiment scores should be interpreted as exploratory results rather than as psychometrically equivalent to standard embodiment scales. Intercoder reliability for sentiment labeling was substantial (kappa = 0.75), supporting the consistency of the sentiment codes. However, reliability was assessed on only a subset of interviews; future work should include additional coders and evaluate consistency in embodiment domains. Additionally, the development of validated instruments to measure embodiment of bionic prostheses, incorporating clinically appropriate control items and established response formats, is an area worth exploring in future studies. As advances in prosthetic technology elicit novel embodiment experiences, tools that are clinically useful and scientifically appropriate for identifying meaningful within- and between-study changes in embodiment are increasingly needed.

Further work may benefit from a take-home component with quantified real-world wear time (*38*) to determine if observed functional and perceptual improvements correlate with sustained real-world adoption (*6*). The interplay between surgery and training remains complex despite the intent of this study to separate their effects. Attribution to surgical revision is strongest for data collected near the beginning of the post-operative testing week, including residual muscle activation, free space reaching performance, and baseline questionnaires regarding the prescribed prosthesis. ROM measures collected at both the beginning and end of each testing week helped distinguish baseline post-surgical changes from within-week training-associated changes. fMRI outcomes were collected at the end of each testing week and therefore reflect the combined influence of surgical revision and training, though the near-term quantity of training is consistent at both timepoints. Subjective bionic embodiment and sentiment measures were administered at the start and end of training to assess exposure-related perceptual changes, but these outcomes are also influenced by prior experience and evolving expectations. A longitudinal study with increased cohort size and additional time points after cessation of training would better separate short-term learning from longer-term neuromechanical changes.

These preliminary findings suggest the rehabilitative potential of the revisional AMI surgery and sustained efferent prosthetic control for embodiment. Near-term adoption of the revisional AMI into the current standard of care might reasonably emphasize home-based therapeutic applications, and may be accelerated through scalable surgical training programs and the availability of commercial prosthetic devices with efferent control capabilities.

## MATERIALS AND METHODS

### Study design

This non-randomized pilot study was designed to: (i) identify potential causal factors for previously observed improvements in neuromuscular function correlated with the AMI (*24–28*); and (ii) characterize the influence of the AMI on subsequent prosthesis embodiment of an efferently controlled bionic knee prosthesis. We formulated a repeated-measures design wherein the intervention is a surgical procedure that revises conventional residual neuromusculature into a revisional AMI for prosthetic knee control. We first hypothesized that the antagonistic architecture of the AMI improves neuromuscular control in the absence of prosthetic training effects. We further hypothesized that the revisional AMI improves prosthetic control and embodiment relative to the levels provided by conventional amputation neuromusculature when controlling for exposure to prosthetic training. Two individuals with preexisting conventional transfemoral amputation were recruited into the study, and an identical set of experiments with increasing exposure to bionic prosthesis control was conducted before and after the surgical intervention. Each set of experiments was administered over the course of seven days to quantify peripheral neuromuscular signaling, central neuromotor activity, and efferent prosthetic control with and without a physical device. Simultaneously, questionnaires and interviews were administered to quantify and qualify subjective prosthesis embodiment throughout each experiment week. An intermediate six month period between experiment weeks allowed participants time to recover from the surgical intervention while facilitating attenuation of training effects.

### Participant recruitment

Two individuals with preexisting conventional transfemoral amputation were recruited for this pilot study. Informed consent for the surgical intervention was provided at Brigham and Women’s Hospital (BWH) (Boston, MA) under the approval of the Partner’s Health System Institutional Review Board (Partners #2019P001681), and as part of the registered clinical trial NCT04063592. Clinical assessments and fMRI experiments were performed at BWH. All other experiments were performed at the MIT Media Lab (Cambridge, MA). The embodiment questionnaires and interviews were approved under the MIT Committee on the Use of Humans as Experimental Subjects protocol #2002000096. Relevant participant biometrics were recorded for subsequent analyses (Table S1). The primary study endpoint was the change in peripheral neuromuscular function from the pre-operative baseline to the six-month post-operative assessment. Secondary endpoints included changes in cortical activity, prosthetic control, embodiment, and proprioception. No adverse events occurred during the study.

### Efferently controlled bionic knee prosthesis

A single degree-of-freedom active knee prosthesis was adapted from the series-elastic design of Carney et al. (*39*) for this study. The prosthesis features a universal titanium pyramid for socket attachment, a passive carbon fiber ankle (AllPro, Fillauer), and a height-adjustable aluminum pylon. The ROM of the knee was limited by hardware and software to 90 degrees of flexion. A serial interface enabled receipt of EMG data streams from custom EMG processing hardware previously designed by our group (*40*). Force sensitive resistors were fixed to the sole of the foot cosmesis at toe, midsole, and heel positions to determine binary ground contact for differentiating between stance and swing phases. Design and control details are reported in our previous work (*28*).

### Statistical Methods

Significance testing for linear regressions was performed using the t-test for coefficients against the null hypothesis that the slope coefficient is 0 (α = 0.05). The two-tailed Welch’s t-test was used to determine significant repeated-participant differences regarding accuracy of free space reaching movements, symmetry of sit-to-stand GRFs, and subjective embodiment of prosthetic devices (α = 0.05). Normality of the data were verified using the Shapiro–Wilk test (α = 0.05). The number of samples to achieve a statistical power of 0.80 (1-β) at a significance level of α = 0.05 in assessments of prosthetic control performance was determined a priori with an estimated effect size of 0.8 (large, detectable by visual inspection). The estimated effect size was informed by the outcomes of previous studies comparing the prosthetic control performance of individuals with the AMI against individuals with conventional amputations (*27*, *28*), in addition to the researchers’ years of collective experience working with the AMI population. Given a repeated-measures experiment design with pre- and post- measurements forming matched pairs, the minimum required sample size per significance test was calculated to be 15 (*41*). Statistical analyses were performed using MATLAB R2023b (The MathWorks Inc.).

## Data Availability

All data associated with this study are available in the paper, the Supplementary Materials, and the public code repositories listed below. All materials used or generated in this study are commercially available or will be provided by the corresponding author upon reasonable request.

## Acknowledgments

We extend our deep gratitude to our clinical collaborators Dr. R. O’Donnell, K. Clites, L. Berger, and R. Chiao. We thank E. Turnator for support with the qualitative analysis. We also thank I. Hayes, J. Cronin, and N. Goderdzishvili for their artistic contributions.

## Author Contributions

T.S., F.R., and H.H. conceptualized the study. T.S., J.M., F.R., J.Q., C.L., Y.T., L.R., M.J.C., C.S., D.L., S.H.Y., E.C., M.N., M.C., and H.H. developed the study methodology, including the surgical, clinical, neurophysiological, imaging, prosthesis, and subjective-assessment approaches. T.S., J.M., F.R., J.Q., C.L., L.R., C.L.S., G.W., P.M., and C.S. performed the investigation and collected experimental data. T.S., J.M., and C.S. performed data visualization and prepared figures. H.H. acquired funding and supervised the project. T.S. and H.H. administered the project. T.S. and J.M. wrote the original draft. T.S., J.M., F.R., J.Q., C.L., Y.T., L.R., M.J.C., C.L.S., G.W., P.M., C.S., D.L., S.H.Y., E.C., M.N., M.C., and H.H. reviewed and edited the manuscript.

